# Asymptomatic SARS-CoV-2 testing: predictors of effectiveness; risk of increasing transmission

**DOI:** 10.1101/2020.11.24.20236950

**Authors:** Jordan P. Skittrall

## Abstract

Testing asymptomatic people for SARS-CoV-2 aims to reduce COVID-19 transmission. Screening programs’ effectiveness depends upon testing strategy, sample handling logistics, test sensitivity, and individual behavior, in addition to dynamics of viral transmission. We investigated the interaction between these factors to determine how to optimize reduction of transmission. We show that under idealistic assumptions 70% of transmission may be averted, but under realistic assumptions only 7% may be averted. We show that programs that overwhelm laboratory capacity or reduce isolation of those with minor symptoms have increased transmission compared with those that do not: programs need to be designed to avoid these issues. Our model allows optimal selection of whom to test, quantifies the balance between accuracy and timeliness, and quantifies potential impacts of behavioral interventions.

**One Sentence Summary:** Programs that overwhelm laboratory capacity or reduce isolation of those with minor symptoms have impaired effectiveness.

## Main Text

Repeatedly screening asymptomatic individuals for SARS-CoV-2, with the aim of isolating infected people and thereby reducing transmission, has been undertaken in hospitals (*1*), institutions (*2, 3*), professional sports leagues (*4*), and the White House. It has been undertaken at town and city level (*5, 6*), with the aim of expansion to national level (*7*). This reduction in transmission aims not only to save lives, but also to permit continuance of activities that would otherwise be halted as part of disease control efforts.

The success of such screening in reducing infections does not rely only upon screening frequency, test sensitivity and viral shedding profiles. It encompasses every element of the screening process, from those affecting whether people with infection undergo screening, through the speed with which screening can inform people they are infectious, to the actions people take on learning they are infectious. Understanding the contributions from and interactions between these elements is key to designing an effective screening program. Crucially, it is key for avoiding a program that loses effectiveness or even increases infection rates by allowing the wrong circumstances to come together.

We have derived an expression for the proportion of infections averted by a screening program (see Supplementary Methods). Our expression accounts for real-world engagement with screening and the time taken to process samples. We show that in realistic situations, screening (with isolation following a positive screen) alone results in only a modest reduction in infections. When the presence of screening results in a relaxation of precautions taken by those with minor symptoms, we show that this combination can result in an overall increase in infections compared with no screening. We demonstrate that the success of screening depends upon a rapid turnaround of tests. As a result, we show that a screening program running comfortably under capacity is more successful than one that pushes capacity and generates backlogs. Our derived expression can be used to compare the effectiveness of proposed testing strategies in complex scenarios where there are different infection rates and different test availabilities – such situations might occur in a small screening program, for example in a hospital with ward-to-ward variation in infection, or in a large program, for example in a national program with city-to-city variation in infection and in laboratory locations.

We began by considering two different screening scenarios. The first scenario is an ideal (maximum impact) scenario. In this scenario, screening is performed daily with a high-sensitivity test. Test turnaround is rapid. All those eligible for screening present on every occasion, and all those with positive screens immediately isolate. The second scenario is a realistic scenario for mass population screening. In this scenario, screening is performed weekly with a high-sensitivity test. Samples for testing must be transported by courier from the sampling site to the testing laboratory. Test turnaround follows that of a large laboratory operating within capacity. There is attrition reducing those eligible who present for screening and those who isolate following positive screens, similar to the attrition observed in other screening programs. The difference between the two scenarios is marked. In the ideal scenario, we estimate screening alone can eliminate 70% of onward transmissions. In the realistic scenario, the proportion of transmissions eliminated reduces to 7%.

We next considered what happens if the presence of screening reassures those with minor symptoms, so that instead of isolating they continue with their daily lives. We divided our infected population into three categories: those who never display any symptoms of infection and always continue with their daily lives, those who display typical symptoms and isolate as soon as these symptoms manifest, and those who display minor symptoms and variably reduce their contact with others when such symptoms manifest. If, instead of isolating, those with minor symptoms behave as usual, the total number of transmissions increases, and because on average an infected person remains infectious for longer (having not isolated), the proportion of transmissions eliminated by screening increases. However, because screening does not identify all the additional individuals in the population with minor symptoms but behaving as usual, the net result is a relative increase in the number of transmissions. If the behavior change is seen in a sufficiently high proportion of those with minor symptoms, the net result of the screening program can be an absolute increase in transmissions compared with no screening program, even though the program appears to be more successful (Fig. 1).

**Fig. 1.**
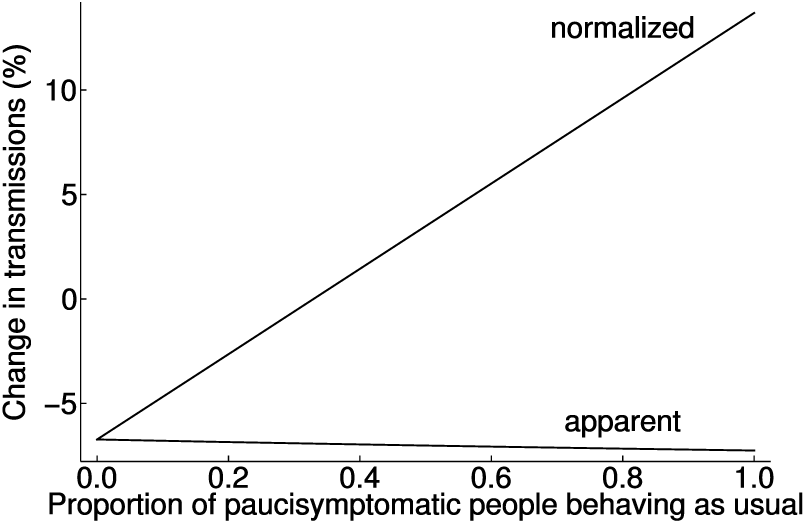
Behavior changes in people with minor symptoms may negate the effect of screening.

Model output where the proportion of those developing minor symptoms (“paucisymptomatic people”) behaving as usual, rather than isolating, is varied from 0 to 1, with parameters otherwise as in our realistic scenario (see Supplementary Methods). As fewer people with minor symptoms isolate, screening detects and leads to isolation of more infectious people (apparent change in transmissions line, showing estimated change in transmissions varying from -6.7% when all paucisymptomatic people isolate, to -7.3% when all paucisymptomatic people behave as usual). However, when the percentage change in transmissions is normalized to a fixed denominator (the number of transmissions without screening when paucisymptomatic people all isolate), it can be seen that the combined effect of screening and behavior change is at best a reduced effectiveness in screening, and at worst an increase in the number of transmissions (normalized change in transmissions line, showing estimated change in transmissions varying from -6.7% when all paucisymptomatic people isolate, to +13.7% when all paucisymptomatic people behave as usual).

Following this, we considered the impact of testing turnaround times on the ability of screening to reduce viral transmission. In general, one would expect a greater number of screening tests to be able to detect a greater number of infections, and therefore to yield a greater reduction in transmissions. However, when the number of tests requested exceeds a laboratory’s capacity, a backlog develops with consequent increase in turnaround time. This effect was seen in English laboratories at the end of April 2020, when a policy of testing large numbers of asymptomatic people in residential facilities was implemented. We modeled the effect of exceeding laboratory capacity using turnaround time data from April–June 2020 in our regional clinical microbiology and public health laboratory in Cambridge, England (Fig. 2) (see Supplementary Methods). Our output shows that reliably keeping laboratory demand slightly below capacity results in a greater reduction in transmissions than when capacity is exceeded. We then proceeded to consider the general effect of laboratory turnaround time on the ability of screening to reduce viral transmission. Our output shows that the extent of transmission reduction depends strongly upon turnaround time.

**Fig. 2.**
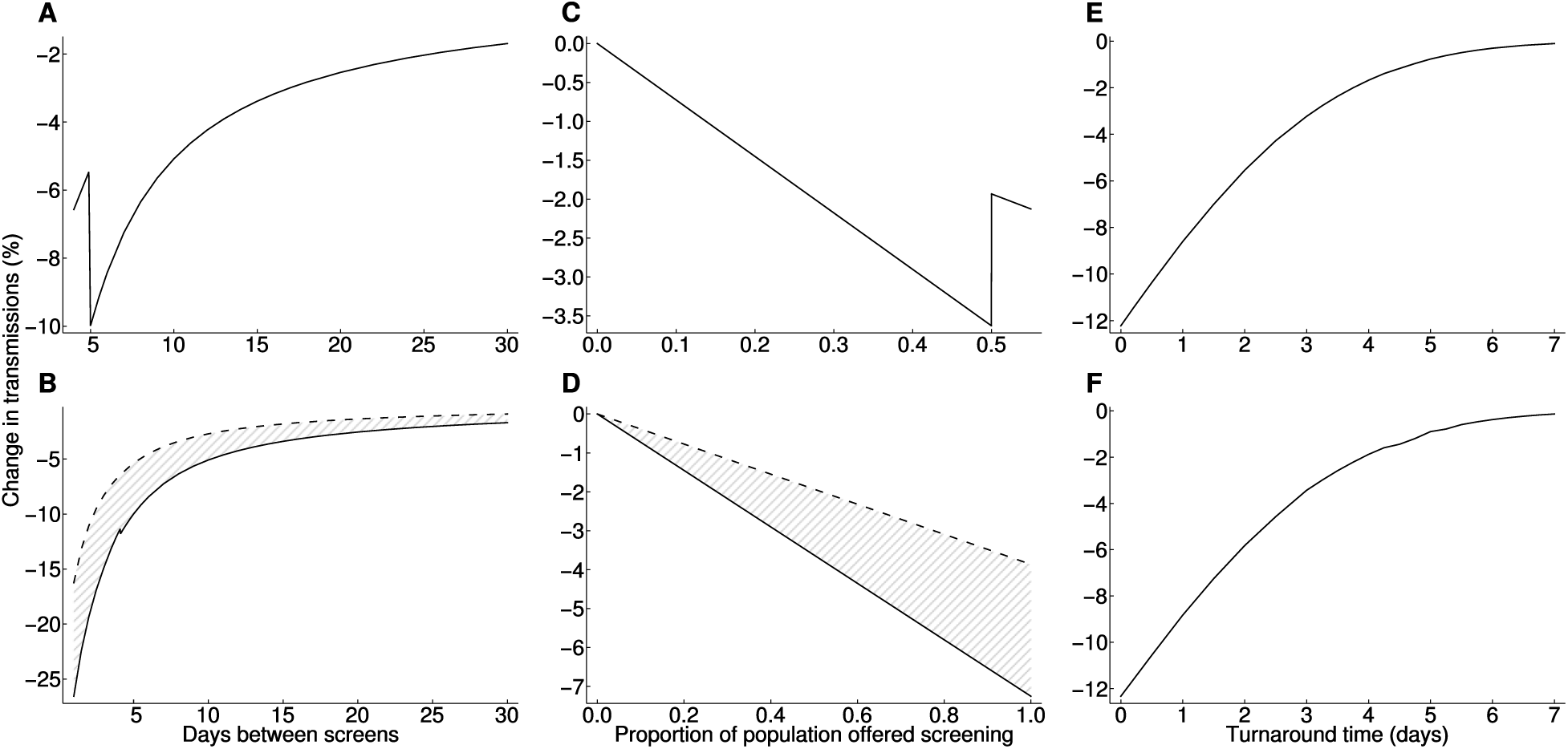
Turnaround time strongly impacts the success of screening. (**A**) Impact on transmissions of shortening the interval between screening tests, until, at a 5-day interval, testing capacity is overwhelmed with an impact on turnaround time. (**B**) Comparison (hatched region) between the effects of normal (solid line) and impaired (dashed line) laboratory turnaround times (see Supplementary Methods) on transmission reduction, for varying screening intervals. (**C**) Impact on transmissions of offering weekly screening to an increasing population proportion until, at 50%, testing capacity is overwhelmed with an impact on turnaround time. (**D**) Comparison (hatched region) between the effects of normal (solid line) and impaired (dashed line) laboratory turnaround times on transmission reduction, for varying proportions of the population offered screening. (**E**) Impact of turnaround time on transmission reduction. Here, rather than being a distribution, total turnaround time from sampling to action on a positive result takes a single value, which is varied. (**F**) As (E), but using reported RNA detection rates from the literature (*8*) rather than assuming the probability of detection scales with infectiousness. This shows the results are not an artefact of assuming detecting infection is more likely in more infectious individuals. Our realistic model parameters are used where not otherwise stated.

A full consideration of screening effectiveness takes into account individual engagement with screening. We considered this in our model, via the proportion of those offered screening who ever take it up, the proportion of those who attend each screening event, and the proportion who isolate when asked. The first and last of these have a linear effect on testing effectiveness (Fig. S1). For a weekly screening interval, the second is also approximately linear, reflecting that this screening interval only gives one opportunity to prevent most transmission. With more frequent screening intervals, the impact on transmissions of increased per-screen uptake becomes nonlinear, with diminishing returns as the uptake is higher (Fig. 3).

**Fig. 3.**
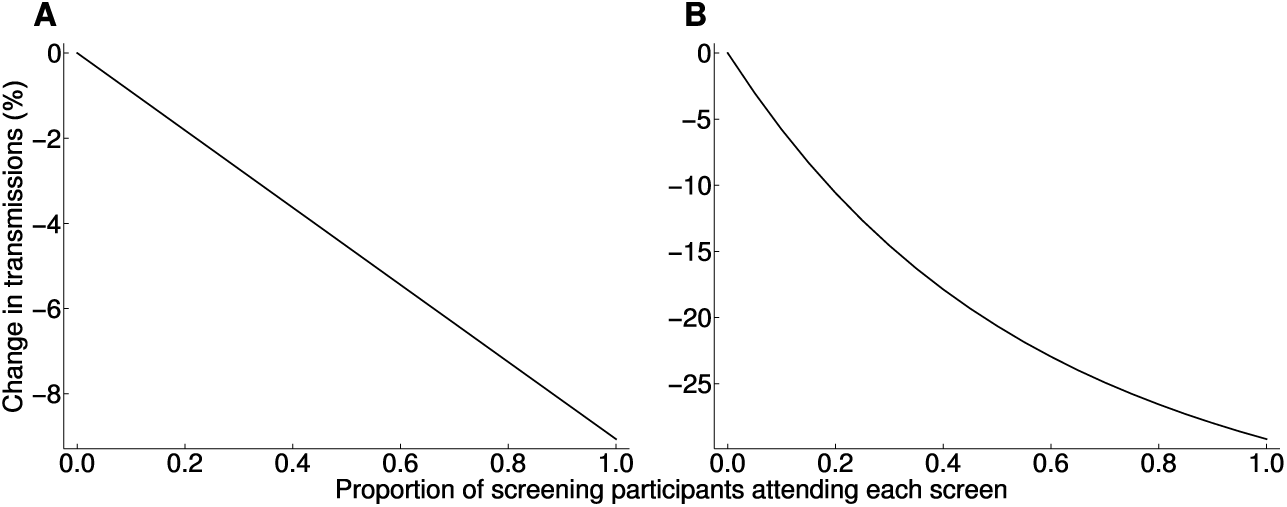
Diminishing returns with increased uptake for short screening intervals. Model output showing change in transmissions as the proportion of those who attend each offered screen (from those who engage in screening at least once) changes. (**A**) Weekly screening interval. (**B**) Daily screening interval. Model parameters are set as for our realistic scenario, except for the proportions attending each screening, and the screening interval in (B).

Because our model takes infectivity and testing distributions as input, limited only by the tractability of the resultant numerical integration, it can be used to predict the impact of different testing strategies in complex scenarios where rates of infection, access to testing or uptake vary within a population (Fig. 4). This allows us to, for example, consider the impact of a program using rapid near-patient tests daily, where such tests have lower sensitivity than laboratory-based nucleic acid amplification testing (*9*). Assuming total and per-test uptake remains the same (increased frequency balanced by increased convenience), our model predicts a reduction in transmissions from such a program between 25% and 32%, compared with the 7% reduction from a centralized mass testing program.

**Fig. 4.**
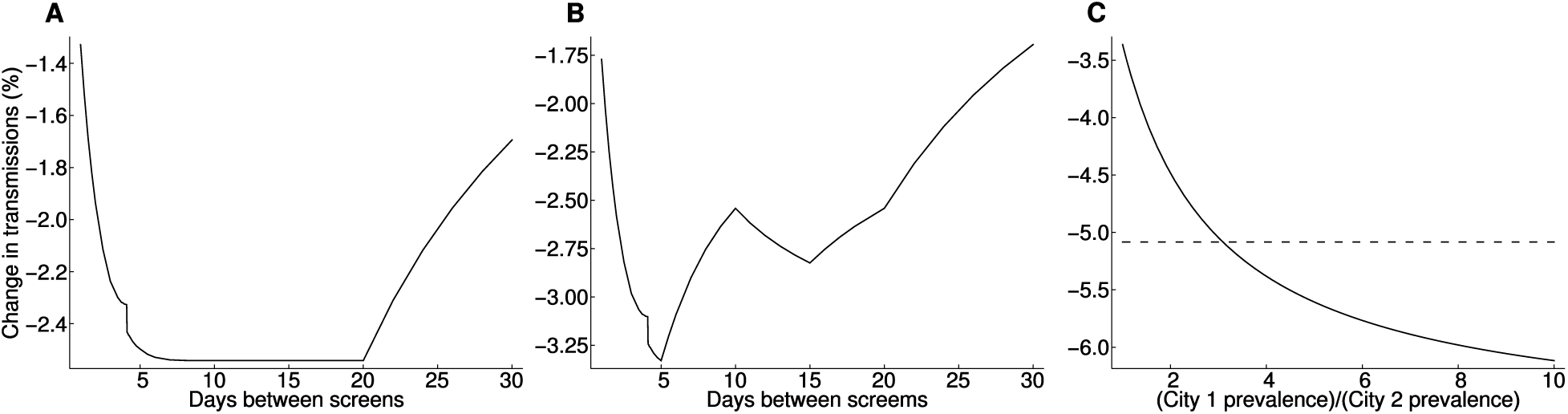
Screening in more complex scenarios. **(A)** Scenario where testing capacity is sufficient to offer screening to the whole population every 20 days (taking non-attendance into account), so that a smaller proportion of the population is screened if testing is offered more frequently. The proportion/frequency combination does not affect testing impact, unless testing is underutilized (right side) or redundant (left side of plot).**(B)** As (A), but with the population structured: half the population are healthcare workers in contact with vulnerable patients, so are always screened before others. Each half of the population further divides into two, with the infection rate in one quarter double that of the other. Those with the higher infection rate are screened first, subject to capacity and the vulnerable patient contact-first rule, i.e. the screening priority is higher rate healthcare workers, lower rate healthcare workers, higher rate others, lower rate others. (**C**) Scenario in which there are two identical cities with two identical laboratories (realistic testing scenario), save that the first city has an infection prevalence greater than the second city. Laboratory capacity is sufficient to offer screening to everybody every 10 days. Each city’s laboratory can be used separately to offer screening to its local population (dashed line). Alternatively, both laboratories can be used to screen every 5 days those in the city with higher infection prevalence, but with an additional two-day turnaround delay for samples sent between cities (solid line). The more effective strategy depends upon relative infection rates in the two cities.

Our model is sufficiently robust to changes in modeling assumptions regarding viral transmission dynamics to make predictions that can be used in practice (Table S1).

Ultimate control of the COVID-19 pandemic is unlikely prior to deployment of effective vaccines. Screening and isolation acts as a bridge to this goal, saving lives and permitting some resumption of economic and social activity. It must, however, be recognized that unintended behavioral responses to screening may not only remove the opportunity to resume desired activity, but indeed may make disease transmission worse than if no screening had occurred.

Pursuing centralized testing strategies may rapidly increase testing capacity. However, the critical impact of turnaround time on testing effectiveness means that such strategies must have highly streamlined logistics chains to retain effectiveness. If rapid centralized turnaround of tests is not possible, then less accurate, localized testing may be more effective. As with the historical introduction of screening programs, in SARS-CoV-2 testing it is crucial we move from simply counting numbers of tests, to more sophisticated measures of their effectiveness.

Engagement with screening substantially impacts success and must not be taken for granted (*10*). A holistic approach considering the social, economic and political impacts, acknowledging the incidence of false positive results and balancing their impact, and combined with good communication, is therefore a key part of a successful program (*11, 12*). Testing strategies that employ confirmatory testing to reduce false positive rates need clear protocols for communicating with those awaiting confirmatory testing, to retain the benefit of earlier isolation.

Our results can only be as accurate as the estimates on which our model is based (for example, of test sensitivity). However, where such estimates limit the accuracy of our predictions, our model highlights the additional information required. (Our model is also limited by numerical accuracy in the calculations, but the underlying estimates represent a greater uncertainty.) In any case, our model demonstrates the impact of the different modifiable factors in a screening program, enabling policymakers to prioritize those that give most benefit.

Our study does not account for the potential impact on transmission rate of isolating contacts of those who screen positive. The impact of contact isolation on transmission reduces as the impact of screening increases (in the extreme case that screening stops all infection, no contact tracing is necessary). As screening identifies more infectious individuals, both the scale of contact tracing required and the impact from isolating non-infectious contacts increase. The relative efficacy of screening is not impacted by contact tracing (although if contact tracing is successful, there should be fewer infectious individuals to screen), so can be considered separately.

Our work allows policymakers to design SARS-CoV-2 screening programs to maximize impact in reducing transmission. Whilst the data used pertain to SARS-CoV-2, the underlying methodology is applicable to any transmissible disease, and can therefore be applied to other epidemics and pandemics.

## Data Availability

Code and novel datasets used to produce results are available from the author. Some evaluated datasets are contained in other publications, as referenced in the text.

## Acknowledgments

I am grateful to colleagues in the Cambridge Clinical Microbiology and Public Health laboratory for their clinical work on SARS-CoV-2 testing, and particularly to data scientist colleagues for assistance obtaining data on turnaround time.

## Supplementary Materials

Materials and Methods Figures S1-S3

Table S1 Code S1

References (*14-38*)

## References and Notes

1. L. Rivett, S. Sridhar, D. Sparkes, M. Routledge, N. K. Jones, S. Forrest, J. Young, J. Pereira-Dias, W. L. Hamilton, M. Ferris, M. E. Torok, L. Meredith, M. Curran, S. Fuller, A. Chaudhry, A. Shaw, R. J. Samworth, J. R. Bradley, G. Dougan, K. G. C. Smith, P. J. Lehner, N. J. Matheson, G. Wright, I. Goodfellow, S. Baker, M. P. Weekes, Screening of healthcare workers for SARS-CoV-2 highlights the role of asymptomatic carriage in COVID-19 transmission. Elife. 9, 1–20 (2020).

2. A. V. Dora, A. Winnett, L. P. Jatt, K. Davar, M. Watanabe, L. Sohn, H. S. Kern, C. J. Graber, M. B. Goetz, Universal and Serial Laboratory Testing for SARS-CoV-2 at a Long-Term Care Skilled Nursing Facility for Veterans — Los Angeles, California, 2020. MMWR. Morb. Mortal. Wkly. Rep. 69, 651–655 (2020).

3. H. Njuguna, M. Wallace, S. Simonson, F. A. Tobolowsky, A. E. James, K. Bordelon, R. Fukunaga, J. A. W. Gold, J. Wortham, T. Sokol, D. Haydel, H. Tran, K. Kim, K. A. Fisher, M. Marlow, J. E. Tate, R. H. Doshi, K. G. Curran, Serial Laboratory Testing for SARS-CoV-2 Infection Among Incarcerated and Detained Persons in a Correctional and Detention Facility - Louisiana, April-May 2020. MMWR. Morb. Mortal. Wkly. Rep. 69, 836–840 (2020).

4. S. M. Kissler, J. R. Fauver, C. Mack, C. Tai, K. Y. Shiue, C. C. Kalinich, S. Jednak, I. M. Ott, C. B. F. Vogels, J. Wohlgemuth, J. Weisberger, J. DiFiori, D. J. Anderson, J. Mancell, D. D. Ho, N. D. Grubaugh, Y. H. Grad, Viral dynamics of SARS-CoV-2 infection and the predictive value of repeat testing. medRxiv (2020), doi:10.1101/2020.10.21.20217042.

5. E. Lavezzo, E. Franchin, C. Ciavarella, G. Cuomo-Dannenburg, L. Barzon, C. Del Vecchio, L. Rossi, R. Manganelli, A. Loregian, N. Navarin, D. Abate, M. Sciro, S. Merigliano, E. De Canale, M. C. Vanuzzo, V. Besutti, F. Saluzzo, F. Onelia, M. Pacenti, S. Parisi, G. Carretta, D. Donato, L. Flor, S. Cocchio, G. Masi, A. Sperduti, L. Cattarino, R. Salvador, M. Nicoletti, F. Caldart, G. Castelli, E. Nieddu, B. Labella, L. Fava, M. Drigo, K. A. M. Gaythorpe, K. E. C. Ainslie, M. Baguelin, S. Bhatt, A. Boonyasiri, O. Boyd, L. Cattarino, C. Ciavarella, H. L. Coupland, Z. Cucunubá, G. Cuomo-Dannenburg, B. A. Djafaara, C. A. Donnelly, I. Dorigatti, S. L. van Elsland, R. FitzJohn, S. Flaxman, K. A. M. Gaythorpe, W. D. Green, T. Hallett, A. Hamlet, D. Haw, N. Imai, B. Jeffrey, E. Knock, D. J. Laydon, T. Mellan, S. Mishra, G. Nedjati-Gilani, P. Nouvellet, L. C. Okell, K. V. Parag, S. Riley, H. A. Thompson, H. J. T. Unwin, R. Verity, M. A. C. Vollmer, P. G. T. Walker, C. E. Walters, H. Wang, Y. Wang, O. J. Watson, C. Whittaker, L. K. Whittles, X. Xi, N. M. Ferguson, A. R. Brazzale, S. Toppo, M. Trevisan, V. Baldo, C. A. Donnelly, N. M. Ferguson, I. Dorigatti, A. Crisanti, Suppression of a SARS-CoV-2 outbreak in the Italian municipality of Vo’. Nature. 584, 425–429 (2020).

6. G. Iacobucci, Covid-19: Mass population testing is rolled out in Liverpool. BMJ. 371, m4268 (2020).

7. G. Iacobucci, R. Coombes, Covid-19?: Government plans to spend £100bn on expanding testing to 10 million a day. BMJ. 370, m3520 (2020).

8. J. Zhao, Q. Yuan, H. Wang, W. Liu, X. Liao, Y. Su, X. Wang, J. Yuan, T. Li, J. Li, S. Qian, C. Hong, F. Wang, Y. Liu, Z. Wang, Q. He, Z. Li, B. He, T. Zhang, Y. Fu, S. Ge, L. Liu, J. Zhang, N. Xia, Z. Zhang, Antibody responses to SARS-CoV-2 in patients of novel coronavirus disease 2019. Clin. Infect. Dis. (2020), doi:10.1093/cid/ciaa344.

9. PHE Porton Down & University of Oxford SARS-CoV-2 LFD test development and validation cell, Preliminary report from the Joint PHE Porton Down & University of Oxford SARS-CoV-2 test development and validation cell: Rapid evaluation of Lateral Flow Viral Antigen detection devices (LFDs) for mass community testing, (available at https://www.ox.ac.uk/sites/files/oxford/media_wysiwyg/UKevaluation_PHEPortonDownUniversityofOxford_final.pdf).

10. L. E. Smith, H. W. W. Potts, R. Amlȏt, N. T. Fear, S. Michie, J. Rubin, Adherence to the test, trace and isolate system: results from a time series of 21 nationally representative surveys in the UK (the COVID-19 Rapid Survey of Adherence to Interventions and Responses [CORSAIR] study). medRxiv (2020), doi:10.1101/2020.09.15.20191957.

11. J. P. Skittrall, M. Wilson, A. A. Smielewska, S. Parmar, M. D. Fortune, D. Sparkes, M. D. Curran, H. Zhang, H. Jalal, Specificity and positive predictive value of SARS-CoV-2 nucleic acid amplification testing in a low prevalence setting. Clin. Microbiol. Infect. (2020), doi:10.1016/j.cmi.2020.10.003.

12. J. P. Skittrall, M. D. Fortune, H. Jalal, H. Zhang, D. A. Enoch, N. M. Brown, A. Swift, Diagnostic tool or screening programme? Asymptomatic testing for SARS-CoV-2 needs clear goals and protocols. Lancet Reg. Heal. - Eur. (2020).

13. Wolfram Research Inc., Mathematica (2020).

14. Q. Zhao, N. Ju, S. Bacallado, R. D. Shah, BETS: The dangers of selection bias in early analyses of the coronavirus disease (COVID-19) pandemic. Ann. Appl. Stat. (2020).

15. W. Xia, J. Liao, C. Li, Y. Li, X. Qian, X. Sun, H. Xu, G. Mahai, X. Zhao, L. Shi, J. Liu, L. Yu, M. Wang, Q. Wang, A. Namat, Y. Li, J. Qu, Q. Liu, X. Lin, S. Cao, S. Huan, J. Xiao, F. Ruan, H. Wang, Q. Xu, X. Ding, X. Fang, F. Qiu, J. Ma, Y. Zhang, A. Wang, Y. Xing, S. Xu, Transmission of corona virus disease 2019 during the incubation period may lead to a quarantine loophole. medRxiv (2020), doi:10.1101/2020.03.06.20031955.

16. S. A. Lauer, K. H. Grantz, Q. Bi, F. K. Jones, Q. Zheng, H. R. Meredith, A. S. Azman, N. G. Reich, J. Lessler, The incubation period of coronavirus disease 2019 (CoVID-19) from publicly reported confirmed cases: estimation and application. Ann. Intern. Med. 172, 577–582 (2020).

17. Z. Hu, C. Song, C. Xu, G. Jin, Y. Chen, X. Xu, H. Ma, W. Chen, Y. Lin, Y. Zheng, J. Wang, Z. Hu, Y. Yi, H. Shen, Clinical characteristics of 24 asymptomatic infections with COVID-19 screened among close contacts in Nanjing, China. Sci. China Life Sci. 63, 706–711 (2020).

18. M. M. Arons, K. M. Hatfield, S. C. Reddy, A. Kimball, A. James, J. R. Jacobs, J. Taylor, K. Spicer, A. C. Bardossy, L. P. Oakley, S. Tanwar, J. W. Dyal, J. Harney, Z. Chisty, J. M. Bell, M. Methner, P. Paul, C. M. Carlson, H. P. McLaughlin, N. Thornburg, S. Tong, A. Tamin, Y. Tao, A. Uehara, J. Harcourt, S. Clark, C. Brostrom-Smith, L. C. Page, M. Kay, J. Lewis, P. Montgomery, N. D. Stone, T. A. Clark, M. A. Honein, J. S. Duchin, J. A. Jernigan, Presymptomatic SARS-CoV-2 Infections and Transmission in a Skilled Nursing Facility. N. Engl. J. Med. 382, 2081–2090 (2020).

19. D. A. Collier, S. M. Assennato, B. Warne, N. Sithole, K. Sharrocks, A. Ritchie, P. Ravji, M. Routledge, D. Sparkes, J. Skittrall, A. Smielewska, I. Ramsey, N. Goel, M. Curran, D. Enoch, R. Tassell, M. Lineham, D. Vaghela, C. Leong, H. P. Mok, J. Bradley, K. G. C. Smith, V. Mendoza, N. Demiris, M. Besser, G. Dougan, P. J. Lehner, M. J. Siedner, H. Zhang, C. S. Waddington, H. Lee, R. K. Gupta, The CITIID-NIHR COVID BioResource Collaboration, Point of Care Nucleic Acid Testing for SARS-CoV-2 in Hospitalized Patients: A Clinical Validation Trial and Implementation Study. Cell Reports Med. 1, 100062 (2020).

20. L. Ferretti, C. Wymant, M. Kendall, L. Zhao, A. Nurtay, L. Abeler-Dörner, M. Parker, D. Bonsall, C. Fraser, Quantifying SARS-CoV-2 transmission suggests epidemic control with digital contact tracing. Science. 368, eabb6936 (2020).

21. S. T. Ali, L. Wang, E. H. Y. Lau, X. K. Xu, Z. Du, Y. Wu, G. M. Leung, B. J. Cowling, Serial interval of SARS-CoV-2 was shortened over time by nonpharmaceutical interventions. Science. 369, 1106–1109 (2020).

22. D. P. Weller, C. Campbell, Uptake in cancer screening programmes: A priority in cancer control. Br. J. Cancer. 101, S55–S59 (2009).

23. E. Lahner, E. Dilaghi, C. Prestigiacomo, G. Alessio, L. Marcellini, M. Simmaco, I. Santino, G. B. Orsi, P. Anibaldi, A. Marcolongo, B. Annibale, C. Napoli, Prevalence of Sars-Cov-2 infection in health workers (HWs) and diagnostic test performance: the experience of a teaching hospital in central Italy. Int. J. Environ. Res. Public Health. 17, 4417 (2020).

24. D. W. Eyre, S. F. Lumley, D. O’Donnell, M. Campbell, E. Sims, E. Lawson, F. Warren, T. James, S. Cox, A. Howarth, G. Doherty, S. B. Hatch, J. Kavanagh, K. K. Chau, P. W. Fowler, J. Swann, D. Volk, F. Yang-Turner, N. E. Stoesser, P. C. Matthews, M. Dudareva, T. Davies, R. H. Shaw, L. Peto, L. O. Downs, A. Vogt, A. Amini, B. C. Young, P. Drennan, A. J. Mentzer, D. Skelly, F. Karpe, M. J. Neville, M. Andersson, A. J. Brent, N. Jones, L. M. Ferreira, T. Christott, B. D. Marsden, S. Hoosdally, R. Cornall, D. W. Crook, D. I. Stuart, G. Screaton, T. E. A. Peto, B. Holthof, A.-M. O’Donnell, D. Ebner, C. P. Conlon, K. Jeffery, T. M. Walker, Differential occupational risks to healthcare workers from SARS-CoV-2 observed during a prospective observational study. Elife. 9, e60675 (2020).

25. A. J. Saah, D. R. Hoover, “Sensitivity” and “specificity” reconsidered: The meaning of these terms in analytical and diagnostic settings. Ann. Intern. Med. 126, 91–94 (1997).

26. Y. Yang, M. Yang, C. Shen, F. Wang, J. Yuan, J. Li, M. Zhang, Z. Wang, L. Xing, J. Wei, L. Peng, G. Wong, H. Zheng, M. Liao, K. Feng, J. Li, Q. Yang, J. Zhao, Z. Zhang, L. Liu, Y. Liu, Evaluating the accuracy of different respiratory specimens in the laboratory diagnosis and monitoring the viral shedding of 2019-nCoV infections. medRxiv (2020), doi:10.1101/2020.02.11.20021493.

27. I. Arevalo-Rodriguez, D. Buitrago-Garcia, D. Simancas-Racines, P. Zambrano-Achig, R. del Campo, A. Ciapponi, O. Sued, L. Martinez-Garcia, A. Rutjes, N. Low, P. Bossuyt, J. Perez-Molina, J. Zamora, False-negative results of initial RT-PCR assays for COVID-19: a systematic review. medRxiv (2020), doi:10.1101/2020.04.16.20066787.

28. S. Woloshin, N. Patel, A. S. Kesselheim, False negative tests for SARS-CoV-2 infection — challenges and implications. N. Engl. J. Med. 383, e38 (2020).

29. J. Lu, J. Peng, Q. Xiong, Z. Liu, H. Lin, X. Tan, M. Kang, R. Yuan, L. Zeng, P. Zhou, C. Liang, L. Yi, L. du Plessis, T. Song, W. Ma, J. Sun, O. G. Pybus, C. Ke, Clinical, immunological and virological characterization of COVID-19 patients that test re-positive for SARS-CoV-2 by RT-PCR. EBioMedicine. 59, 102960 (2020).

30. Y. Sohn, S. J. Jeong, W. S. Chung, J. H. Hyun, Y. J. Baek, Y. Cho, J. H. Kim, J. Y. Ahn, J. Y. Choi, J.-S. Yeom, Assessing Viral Shedding and Infectivity of Asymptomatic or Mildly Symptomatic Patients with COVID-19 in a Later Phase. J. Clin. Med. 9, 2924 (2020).

31. J. Bullard, K. Dust, D. Funk, J. E. Strong, D. Alexander, L. Garnett, C. Boodman, A. Bello, A. Hedley, Z. Schiffman, K. Doan, N. Bastien, Y. Li, P. G. Van Caeseele, G. Poliquin, Predicting infectious Severe Acute Respiratory Syndrome Coronavirus 2 from diagnostic samples. Clin. Infect. Dis., ciaa638 (2020).

32. R. A. P. M. Perera, E. Tso, O. T. Y. Tsang, D. N. C. Tsang, K. Fung, Y. W. Y. Leung, A. W. H. Chin, D. K. W. Chu, S. M. S. Cheng, L. L. M. Poon, V. W. M. Chuang, M. Peiris, SARS-CoV-2 virus culture and subgenomic RNA for respiratory specimens from patients with mild Coronavirus disease. Emerg. Infect. Dis. 26, 2701–2704 (2020).

33. K. Inagaki, M. S. Song, J. C. Crumpton, J. DeBeauchamp, T. Jeevan, E. I. Tuomanen, R. J. Webby, H. Hakim, Correlation between the interval of influenza virus infectivity and results of diagnostic assays in a ferret model. J. Infect. Dis. 213, 407–410 (2016).

34. S. Bauerle Bass, S. B. Ruzek, L. Ward, T. F. Gordon, A. Hanlon, A. J. Hausman, M. Hagen, If you ask them, will they come? Predictors of quarantine compliance during a hypothetical avian influenza pandemic: results from a statewide survey. Disaster Med. Public Health Prep. 4, 135–144 (2010).

35. R. K. Webster, S. K. Brooks, L. E. Smith, L. Woodland, S. Wessely, G. J. Rubin, How to improve adherence with quarantine: rapid review of the evidence. Public Health. 182, 163–169 (2020).

36. M. Bodas, K. Peleg, Self-isolation compliance in the COVID-19 era influenced by compensation: findings from a recent survey in Israel. Health Aff. 39, 936–941 (2020).

37. A. C. Marcus, L. A. Crane, C. P. Kaplan, A. E. Reading, E. Savage, J. Gunning, G. Bernstein, J. S. Berek, Improving adherence to screening follow-up among women with abnormal pap smears: results from a large clinic-based trial of three intervention strategies. Med. Care. 30, 216–230 (1992).

38. C.-S. Kuo, G.-R. Chen, S.-H. Hung, Y.-L. Liu, K.-C. Huang, S.-Y. Cheng, Women with abnormal screening mammography lost to follow-up: an experience from Taiwan. Medicine (Baltimore). 95, e3889 (2016).

