## Supplementary Materials for "Asymptomatic SARS-CoV-2 testing: predictors of effectiveness; risk of increasing transmission"

Jordan P. Skittrall

#### This PDF file includes:

Materials and Methods
Figs. S1 to S3
Table S1
Caption for Code S1

#### Other Supplementary Materials for this manuscript include the following:

Code S1 – available on request from the author

### Materials and Methods

#### Model Derivation

Our model considers the total infectivity of a population over a period of time, and the proportion of this infectivity that is averted by screening and isolating infectious people. Infectivity is deemed to end naturally at the end of asymptomatic viral shedding, or when an individual develops symptoms and isolates from others. We construct the model by starting with a simplified screening scenario. We then add more realistic components stepwise until we have reached the final model. Ultimately, the model takes as input distributions of virus shedding duration, infectivity, intervals between tests, testing turnaround times, detection probabilities, and population engagement with screening.

We begin by considering the scenario represented in Fig. S2. Asymptomatic/presymptomatic shedding lasts for duration $m \in M$. It begins during a screening interval of duration $j \in J$ (in principle, successive screening interval durations can be drawn from different distribtions $J_2, J_3, \ldots$). The shedding begins $k \in K$ time prior to the end of the interval. There is a delay $d \in D$ from the screening event prior to isolating an infectious individual, reflecting the turnaround time of the test.

There is a variable probability of transmission (relative infectivity) during the asymptomatic/presymptomatic shedding period. At its most general, this can be expressed as a normalized force of infection $g(t, \tau)$, satisfying

$$g(t, \tau) = 0,\ t < 0 \text{ or } t > \tau,$$

$$\int_0^\infty \int_0^\tau g(t, \tau) f_M(\tau)\, \mathrm{d}t\, \mathrm{d}\tau = 1.$$

With this definition, in the absence of screening the total normalized infectivity is 1. The total normalized infectivity with screening and isolation is therefore expressed as a proportion. This proportion can be used as a coefficient to determine the effect of screening on the reproduction number $R$. In this paradigm, relative infectivity depends upon both position in the shedding period and total duration of shedding: groups with different shedding durations can have different total infectivities. $g$ can be interpreted as a composite reflecting the average shedding profile of an individual. The most obvious way in which $g$ can be seen to be a composite is that if the same shedding period can represent both asymptomatic and presymptomatic shedding, then $g$ represents a composite of the two normalized by their respective probabilities.

Next, we reflect that screening does not pick up all infectious individuals by introducing a probability of detection $p_d(t, \tau) \in [0, 1]$. This probability is allowed to depend on the total duration of shedding. It represents a composite of the probability that an individual presents for screening, the sensitivity of the screening test, and the probability that the individual isolates following a positive test. For nucleic acid amplification testing for SARS-CoV-2, both $g(t, \tau)$ and $p_d(t, \tau)$ are related to viral shedding, and hence they are related to each other. However, shedding of non-viable RNA later in the course of infection means that $p_d(t, \tau)$ tapers more slowly than $g(t, \tau)$ over time.

We now proceed to derive a generalized expression for the relative infectivity in the presence of a given screening regimen. First, we derive an expression for $f_K(x)$, the density function for the amount of time before the first screen that shedding starts. We assume a constant hazard for the commencement of shedding over time. This assumption is valid as long as screening maintains an equilibrium of infections (there is not a large growth or contraction in infection prevalence between successive screens), and as long as the time of onset of infectiousness can be assumed independent between individuals (true in all but very small populations). Note, however, that the screening interval length $j$ can vary. This means that the time before the first screen that shedding starts, $K$, is not simply distributed uniformly across the sampled screening interval $j$, but needs to be conditioned upon the probability that shedding begins in a particular interval. For $J'$ the distribution of the screening interval length in which the shedding begins, we have

$$f_{J'}(x) \propto x f_J(x),$$

and by integrating both sides with respect to $x$ we can derive the constant of proportionality and conclude that

$$f_{J'}(x) = \frac{x f_J(x)}{\mathbb{E}(J)}.$$

It then follows that

$$\begin{aligned} f_K(x) &= \int_0^\infty \frac{f_{J'}(y)}{y} I(x \in (0, y))\,\mathrm{d}y \\ &= \int_x^\infty \frac{f_J(y)}{\mathbb{E}(J)}\,\mathrm{d}y \\ &= \frac{1 - F_J(x)}{\mathbb{E}(J)}. \end{aligned}$$

Now that we have a density function for the time from the start of shedding to the next screen, we can write down a density function for the time from the start of shedding to the time an individual with a positive screen result can be isolted. This is simply the convolution

$$\begin{aligned} f_{K+D}(x) &= \int_0^\infty f_K(y) f_D(x-y)\,\mathrm{d}y \\ &= \int_0^\infty \frac{1 - F_J(y)}{\mathbb{E}(J)} f_D(x-y)\,\mathrm{d}y. \end{aligned}$$

This convolution is independent of the total shedding duration. However, its sampled value can exceed the total shedding duration, that is, shedding can have ended before it is possible to act on a screening result.

We can now write down our first, simplified expression for averted infectivity. In this simplified case a screen always identifies an infectious individual, and that individual then perfectly isolates. Conditioned upon a total shedding time $\tau$, the normalized infectivity averted is

$$\int_0^\infty f_{K+D}(t) \int_{\min(t,\tau)}^\tau g(x,\tau)\,\mathrm{d}x\,\mathrm{d}t,$$

where the $\min$ reflects the possibility of shedding having ended prior to the screen.

Using $g(x,\tau) = 0$ for $x > \tau$ we can expand the components of the integral to give a normalized infectivity averted for shedding time $\tau$ of

$$A_1(\tau) = \int_0^\tau \int_0^\infty \int_t^\tau \frac{1 - F_J(y)}{\mathbb{E}(J)} f_D(t-y) g(x,\tau)\,\mathrm{d}x\,\mathrm{d}y\,\mathrm{d}t.$$

The total normalized infectivity averted from one infectivity-abolishing screen is therefore

$$\begin{aligned} A_1 &= \int_0^\infty A_1(\tau) f_M(\tau)\,\mathrm{d}\tau \\ &= \int_0^\infty \int_0^\tau \int_0^t \int_t^\tau f_M(\tau) \left(\frac{1 - F_J(y)}{\mathbb{E}(J)}\right) f_D(t-y) g(x,\tau)\,\mathrm{d}x\,\mathrm{d}y\,\mathrm{d}t\,\mathrm{d}\tau. \end{aligned}$$

Next, we consider the situation where the probability that a screen abolishes infectivity need not be $1$. In this case, conditioned upon an overall probability $p_\infty$ of *ever* detecting infection in an individual given arbitrarily many tests over arbitrarily short testing intervals, a successful screen at time $y$ occurs with probability $p(y,\tau)$ and unsuccessful screens happen at time $a_1, a_1 + a_2, a_1 + a_2 + a_3, \ldots < y$ with probabilities $1 - p(a_1,\tau), 1 - p(a_1 + a_2,\tau), 1 - p(a_1 + a_2 + a_3,\tau), \ldots$. (In our earlier notation, $p_d(t,\tau) = p_\infty p(t,\tau)$.) Note that the length of shedding in the interval before the first screen is still described by the random variable $K$, but in subsequent intervals shedding occurs across the entire interval unless it finishes altogether, so is described by $J_i$, $i \geq 2$. Fig. S3 gives a schematic representation of this setup to illustrate the notation used. Testing turnaround time is only applicable after the last, successful, screen. (If turnaround time is highly variable, it is in principle possible to have two positive screens with the result of the second available before the first. In practice, most laboratory testing is first in, first out, so we neglect this possibility.)

For any given number of screens prior to successful detection, we can express the infectivity averted by taking a convolution of the distributions of testing times as an extension to the expression for $f_{K+D}(x)$. This yields a general expression for the distribution of the time from beginning of shedding (with total shedding length $\tau$) to isolation of an individual with a positive test result:

$$
\begin{aligned}
f_T(t|\tau) = p_\infty &\int_0^t p(y,\tau) f_K(y) f_D(t-y)\,\mathrm{d}y \\
&+ p_\infty \int_0^t \int_0^y p(y,\tau)\left(1-p(a_1,\tau)\right) f_K(a_1) f_{J_2}(y-a_1) f_D(t-y)\,\mathrm{d}a_1\,\mathrm{d}y \\
&+ p_\infty \int_0^t \int_0^y \int_0^{y-a_1} p(y,\tau)\left(1-p(a_1,\tau)\right)\left(1-p(a_2,\tau)\right) f_K(a_1) f_{J_2}(a_2) \cdot \\
&\qquad\qquad \cdot f_{J_3}(y-a_1-a_2) f_D(t-y)\,\mathrm{d}a_2\,\mathrm{d}a_1\,\mathrm{d}y + \ldots \\
&+ p_\infty \int_0^t \int_0^y \int_0^{y-a_1} \int_0^{y-a_1-a_2} \cdots \int_0^{y-a_1-\ldots-a_{n-2}} p(y,\tau)\left(1-p(a_1,\tau)\right)\left(1-p(a_1+a_2,\tau)\right)\cdot\ldots \\
&\qquad \ldots\cdot\left(1-p(a_1+a_2+\ldots+a_{n-1},\tau)\right) f_K(a_1) f_{J_2}(a_2) f_{J_3}(a_3)\cdot\ldots \\
&\qquad \ldots\cdot f_{J_{n-1}}(a_{n-1}) f_{J_n}(y-a_1-\ldots-a_{n-1}) f_D(t-y)\,\mathrm{d}a_{n-1}\,\mathrm{d}a_{n-2}\ldots\,\mathrm{d}a_1\,\mathrm{d}y \\
&+ \ldots.
\end{aligned}
$$

The total infectivity averted from screening is therefore

$$
\begin{aligned}
A &= \int_0^\infty \int_0^\infty \int_{\min(t,\tau)}^\tau f_M(\tau) f_T(t|\tau) g(x,\tau)\,\mathrm{d}x\,\mathrm{d}t\,\mathrm{d}\tau \\
&= p_\infty \sum_{n=1}^\infty A_n,
\end{aligned}
$$

where

$$
\begin{aligned}
A_1 &= \int_0^\infty\!\!\int_0^\tau\!\!\int_0^t\!\!\int_t^\tau f_M(\tau) p(y,\tau)\left(\frac{1-F_J(y)}{\mathbb{E}(J)}\right) f_D(t-y) g(x,\tau)\,\mathrm{d}x\,\mathrm{d}y\,\mathrm{d}t\,\mathrm{d}\tau, \\
A_2 &= \int_0^\infty\!\!\int_0^\tau\!\!\int_0^t\!\!\int_0^y\!\!\int_t^\tau f_M(\tau) p(y,\tau)\left(1-p(a_1,\tau)\right)\left(\frac{1-F_J(a_1)}{\mathbb{E}(J)}\right) f_{J_2}(y-a_1)\cdot \\
&\qquad\qquad \cdot f_D(t-y) g(x,\tau)\,\mathrm{d}x\,\mathrm{d}a_1\,\mathrm{d}y\,\mathrm{d}t\,\mathrm{d}\tau, \\
A_n &= \int_0^\infty\!\!\int_0^\tau\!\!\int_0^t\!\!\int_0^y\!\!\int_0^{y-a_1}\!\!\int_0^{y-a_1-a_2}\!\!\cdots\int_0^{y-a_1-\ldots-a_{n-2}}\int_t^\tau f_M(\tau) p(y,\tau)\cdot \\
&\quad \cdot\left(1-p(a_1,\tau)\right)\left(1-p(a_1+a_2,\tau)\right)\ldots\left(1-p(a_1+a_2+\ldots+a_{n-1},\tau)\right)\cdot \\
&\quad \cdot\left(\frac{1-F_J(a_1)}{\mathbb{E}(J)}\right) f_{J_2}(a_2) f_{J_3}(a_3)\ldots f_{J_{n-1}}(a_{n-1}) f_{J_n}(y-a_1-\ldots-a_{n-1})\cdot \\
&\quad \cdot f_D(t-y) g(x,\tau)\,\mathrm{d}x\,\mathrm{d}a_{n-1}\,\mathrm{d}a_{n-2}\ldots\,\mathrm{d}a_1\,\mathrm{d}y\,\mathrm{d}t\,\mathrm{d}\tau.
\end{aligned}
$$

These integrals are high-dimensional, but we have successfully implemented code to calculate them in Mathematica (*13*), using its numerical integration algorithms. (We use the global adaptive numerical integration method unless otherwise stated, but have implemented other methods in the code for cross-checking.) The bounds on the integrals mean that the first few terms in the sum are computationally tractable in full.

When successive screens are close together, so that multiple terms in the sum become relevant, performing the full multidimensional integrals becomes computationally intractable. However, when the time between one screening opportunity and the following opportunity is very similar for all individuals eligible for screening, the above convolutions can be markedly simplified. The appropriate simplification is the approximation $f_{J_i}(t) = \delta(t - t_i)$ for $i \geq 2$, where $\delta$ is the Dirac delta and $t_i$ are fixed intervals between successive screens (which may differ from each other). This allows reduction of all the $A_i$ to four-dimensional integrals: $A_1$ is unchanged, and we have

$$A_2 = \int_{t_2}^{\infty}\int_{t_2}^{\tau}\int_{t_2}^{t}\int_{t}^{\tau} f_M(\tau)p(y,\tau)\left(1 - p(y - t_2,\tau)\right)\left(\frac{1 - F_J(y - t_2)}{\mathbb{E}(J)}\right)\cdot \cdot f_D(t - y)g(x,\tau)\,\mathrm{d}x\,\mathrm{d}y\,\mathrm{d}t\,\mathrm{d}\tau,$$

$$A_n = \int_{t_2+t_3+\ldots+t_n}^{\infty}\int_{t_2+t_3+\ldots+t_n}^{\tau}\int_{t_2+t_3+\ldots+t_n}^{t}\int_{t}^{\tau} f_M(\tau)p(y,\tau)\cdot \cdot\left(1 - p(y - t_2 - t_3 - \ldots - t_n,\tau)\right)\left(1 - p(y - t_3 - \ldots - t_n,\tau)\right)\cdot\ldots\cdot\left(1 - p(y - t_n,\tau)\right)\cdot \cdot\left(\frac{1 - F_J(y - t_2 - t_3 - \ldots - t_n)}{\mathbb{E}(J)}\right) f_D(t - y)g(x,\tau)\,\mathrm{d}x\,\mathrm{d}y\,\mathrm{d}t\,\mathrm{d}\tau.$$

In a program undertaking many screens at short intervals, this approximation is likely to prove more accurate than early truncation of a series containing the full, multiple-integral, terms. We note that the approximation is better as $p(t,\tau)$ is more smooth, although the variability in the time to initial sampling means that even when $p(t,\tau)$ is rapidly varying, the approximation is still good as long as the intervals between successive screens are similar.

#### Model Parameters

To evaluate the effects of different screening scenarios, we set parameters of the model according to estimates for SARS-CoV-2 or the parameters we wished to evaluate.

**Duration of asymptomatic/presymptomatic viral shedding.** The distribution $M$ of the duration of asymptomatic/presymptomatic shedding is a composite of two distributions: $M_P$, the duration of presymptomatic shedding in those who eventually develop symptoms, and $M_A$, the total duration of shedding in those who never develop symptoms sufficient that they isolate.

We estimated the duration of presymptomatic shedding in those who eventually develop symptoms as follows. We began with data from Zhao *et al.* (*14*). This study took 378 individuals representing cases of COVID-19 exported from Wuhan, in order to infer, *inter alia*, a non-parametric incubation period distribution that takes into account a number of other biases not fully considered by other authors. The authors then estimated a parametric distribution for the incubation period: Gamma distribution with median 4.5 days and 95-centile 13.4 days (which can be solved numerically to derive shape and scale parameters of 1.81 and 3.03, respectively). We set the duration of presymptomatic shedding to be this full incubation period. We took into account that an individual may be non-infectious or minimally infectious during some of this time in the transmission probability parameter.

To evaluate the impact of assuming different presymptomatic shedding distributions, we also cross-checked our model using distrubutions inferred from two other publications. Xia *et al.* (*15*) took 106 individuals with short potential contacts (less than three days) either with another individual known to be infected with SARS-CoV-2 or by visiting Wuhan province, China. They inferred asymptomatic periods from the dates of symptom onset, fitting the data to a Weibull distribution with estimated mean and median of 4.9 and 4.5, respectively. Lauer *et al.* (*16*) took 181 individuals representing publicly reported confirmed cases with short windows of exposure and symptom onset. They inferred incubation periods and fitted these to a log-normal model with mean and median of 5.5 and 5.1 days, respectively. (Where not otherwise stated, we calculated presymptomatic shedding using the Gamma distribution derived by Zhao *et al.*)

We estimated the duration of viral shedding in those who never go on to develop symptoms by taking the distribution of shedding of the 19 patients in the study by Hu *et al.* (*17*). This study considered detection of SARS-CoV-2 RNA from pharyngeal swabs by quantitative polymerase chain reaction (qPCR). It did not follow up all patients to the point of serial non-detection of RNA, and so we took the first non-detection of RNA as the point of cessation of viral shedding in deriving the distribution we used, even if there was detection of RNA following this event. The dynamics of asymptomatic shedding of virus by those who never go on to develop symptoms are not well-studied, and this decision is equivalent to estimating that shedding of viable virus ceases the first time no RNA can be detected.

The above steps give the distributions $M_P$ and $M_A$ of presymptomatic and asymptomatic viral shedding duration, respectively. To derive the combined distribution, $M$, requires knowing the relative proportions in a given infectious population of those who are presymptomatic and those who are asymptomatic. We denote the proportion of presymptomatic individuals by $\phi$. This value may vary depending upon the population type. More importantly, however, the value may vary within a single population that experiences the same distribution of symptoms, depending upon behavior of individuals. We therefore considered three categories of individual: truly asymptomatic individuals (those who never develop any symptoms), typically symptomatic individuals (those who develop typical symptoms of the disease, sufficient to cause them to isolate), and paucisymptomatic individuals (those who develop symptoms, but who may or may not isolate, depending upon external factors). The paucisymptomatic individuals were then dichotomized into behaving as asymptomatic or as symptomatic. The proportion continuing normal behavior may depend upon clinical advice but also upon setting and population demographics. In particular, the presence of asymptomatic screening itself may affect the behavior of paucisymptomatic individuals. Although the behavior and overall infectiousness of paucisymptomatic individuals may be intermediate between asymptomatic and symptomatic individuals, at population level it can be modeled by the dichotomy we have described, with appropriate adjustment of infectiousness parameters.

To determine estimates for the possible proportions of individuals who remain truly asymptomatic, or become paucisymptomatic or symptomatic, we considered two publications. Arons *et al.* (*18*) conducted point prevalence surveys in an entire residential facility. Of the 55 individuals with SARS-CoV-2 detections in point prevalence surveys, 3 were always asymptomatic and at least 34 had or developed typical symptoms, yielding edge case symptomatic proportions of 62% and 95%. Rivett *et al.* (*1*) conducted screening of asymptomatic healthcare workers. Of the 19 healthcare workers who had not experienced any symptoms before time of screening, 1 went on to develop symptoms, 5 never had any symptoms, and 13 had atypical symptoms, yielding edge case symptomatic proportions of 5% and 74%. Using either study to estimate proportions of individuals experiencing symptoms incorporates an ascertainment bias (both were undertaken in populations with a relatively high number of symptomatic individuals). The screens were point prevalence surveys, but because the populations being screened were, relatively speaking, closed, especially in the residential facility case, there is likely to be a correlation between the timings of infection in those without and those with symptoms. Our model is therefore calculated to provide agreement with these observed figures taking into account differences in the model estimates for the probability of a successful screen between those with and without symptoms, but not differences in duration of shedding. We note that some of the detection probability models we use do not have any dependence upon presence of symptoms, but where there is an implicit dependence (via the detection probability being proportional to the transmission probability, which is allowed to vary depending upon presence of symptoms), we took this into account via a crude scaling.

Unless otherwise stated, our calculations use proportions of asymptomatic individuals derived from both publications noted above, and class all paucisymptomatic individuals amongst those who are asymptomatic. (That is, we assumed that those with minor symptoms continue to behave as usual.)

**Screening interval.** The screening intervals $J_n$ are in principle set as part of the screening strategy and can be varied to evaluate the impact upon transmission. We evaluated a range of possible fixed screening intervals. In practice, even with a screening interval of a fixed number of days, there may be some variation in time of day when the sample is taken, which for short periods of asymptomatic shedding may prove significant. This could be modeled by considering the differences between successive uniform distributions over single working days of screening. However, we anticipated the loss of accuracy caused by needing higher dimensional numerical integration of the resultant conditional distributions would outweigh the approximation of a fixed interval.

**Test turnaround time.** Test turnaround consists of a number of steps, which divide broadly into the time to transport the sample to a laboratory, time to produce a result in the laboratory, and time from test result until that individual is isolated and unable to infect others. These can vary substantially between screening scenarios. At one extreme is a near-instantaneous near-patient test; at the other extreme is a slow laboratory turnaround with a long logistics chain to deliver samples and results.

To compare effects of laboratory turnaround time, we considered two scenarios, based upon data from the Cambridge Clinical Microbiology and Public Health Laboratory, which tests samples in the East of England:

- Working below capacity, processing samples continuously. The data for this scenario are taken from a seven-day period (samples received 00:00 14$^{\text{th}}$–23:59 20$^{\text{th}}$ May 2020) of recorded laboratory turnaround times.

- Capacity exceeded, processing samples continuously. The data for this scenario are taken from a seven-day period (samples received 00:00 28$^{\text{th}}$ April–23:59 4$^{\text{th}}$ May 2020) of recorded laboratory turnaround times. During the period considered, an increase in screening samples from residential facilities coincided with a change in the platform used in part of the analytical process (to increase capacity and avoid reagent shortages). In theory, when capacity is exceeded the turnaround time would continue to grow. However, in practice the laboratory introduces mitigation measures such as steps to increase capacity, to reduce demand, and to contact other laboratories for assistance with testing. We include this scenario to model the effect of increasing the screening rate beyond the capacity of a laboratory to process samples at optimal turnaround time.

The steps affecting turnaround time outside a laboratory depend substantially upon the scenario. We added to our model two further possible delays to turnaround: a fixed delay representing usually constant factors (such as transport to the laboratory within a hospital or using a courier whose journey time varies little) and a variable delay modelled using a uniform distribution from zero to a given time period, representing a periodical event such as a courier pickup or a human checking results. The delays were added via a convolution of probability distribution functions to derive $f_D$, in the case of the variable delay, and by applying an offset to $f_D$, in the case of the fixed delay. These can be calculated analytically, either by using the laboratory turnaround time datasets as probability mass distributions, or by turning those datasets into histograms and deriving piecewise continuous probability density functions. We chose the latter option for ease of comparison with the case when no additional delay is present.

We also considered scenarios with different, fixed, turnaround times, so that turnaround time could be varied continuously to evaluate its effect. Within these scenarios was a "best achievable" setup, of 90-minute turnaround. This represents the ideal situation where a sample is immediately run on a rapid point-of-care platform and results are available and acted upon immediately. The 90 minute figure is derived from the sample processing time for SARS-CoV-2 testing using the SAMBA II platform (*19*). In this scenario the test is run as soon as the sample is taken and there is no delay in acting on results, so that $f_D(t) = \delta(t - t_{\min})$, where $\delta$ is the Dirac delta function and $t_{\min}$ is the ideal laboratory turnaround time of 90 minutes. The multidimensional integrals may then be reduced to

$$A_1 = \int_{t_{\min}}^{\infty}\int_{t_{\min}}^{\tau}\int_{t}^{\tau} f_M(\tau)p(t - t_{\min}, \tau)\left(\frac{1 - F_J(t - t_{\min})}{\mathbb{E}(J)}\right) g(x, \tau)\,\mathrm{d}x\,\mathrm{d}t\,\mathrm{d}\tau,$$

$$A_2 = \int_{t_{\min}}^{\infty}\int_{t_{\min}}^{\tau}\int_{0}^{t-t_{\min}}\int_{t}^{\tau} f_M(\tau)p(t - t_{\min}, \tau)\,(1 - p(a_1, \tau))\left(\frac{1 - F_J(a_1)}{\mathbb{E}(J)}\right)\cdot$$

$$\cdot\, f_{J_2}(t - t_{\min} - a_1)g(x, \tau)\,\mathrm{d}x\,\mathrm{d}a_1\,\mathrm{d}t\,\mathrm{d}\tau,$$

$$A_n = \int_{t_{\min}}^{\infty}\int_{t_{\min}}^{\tau}\int_{0}^{t-t_{\min}}\int_{0}^{t-t_{\min}-a_1}\int_{0}^{t-t_{\min}-a_1-a_2}\cdots\int_{0}^{t-t_{\min}-a_1-\ldots-a_{n-2}}\int_{t}^{\tau} f_M(\tau)\cdot$$

$$\cdot\, p(t - t_{\min}, \tau)\,(1 - p(a_1, \tau))\,(1 - p(a_1 + a_2, \tau))\ldots(1 - p(a_1 + a_2 + \ldots + a_{n-1}, \tau))\cdot$$

$$\cdot\left(\frac{1 - F_J(a_1)}{\mathbb{E}(J)}\right) f_{J_2}(a_2)f_{J_3}(a_3)\ldots f_{J_{n-1}}(a_{n-1})f_{J_n}(t - t_{\min} - a_1 - \ldots - a_{n-1})\cdot$$

$$\cdot\, g(x, \tau)\,\mathrm{d}x\,\mathrm{d}a_{n-1}\,\mathrm{d}a_{n-2}\ldots\,\mathrm{d}a_1\,\mathrm{d}t\,\mathrm{d}\tau.$$

If we additionally use the approximation of fixed intervals between successive screens, this reduces further: $A_1$ is unchanged, and we have

$$A_2 = \int_{t_{\min}+t_2}^{\infty}\int_{t_{\min}+t_2}^{\tau}\int_{t}^{\tau} f_M(\tau)p(t - t_{\min}, \tau)\,(1 - p(t - t_{\min} - t_2, \tau))\cdot$$

$$\cdot\left(\frac{1 - F_J(t - t_{\min} - t_2)}{\mathbb{E}(J)}\right) g(x, \tau)\,\mathrm{d}x\,\mathrm{d}t\,\mathrm{d}\tau,$$

$$A_n = \int_{t_{\min}+t_2+\ldots+t_n}^{\infty}\int_{t_{\min}+t_2+\ldots+t_n}^{\tau}\int_{t}^{\tau} f_M(\tau)p(t - t_{\min}, \tau)\cdot$$

$$\cdot\,(1 - p(t - t_{\min} - t_2 - t_3 - \ldots - t_n, \tau))\,(1 - p(t - t_{\min} - t_3 - \ldots - t_n, \tau))\cdot\ldots$$

$$\ldots\cdot(1 - p(t - t_{\min} - t_n, \tau))\left(\frac{1 - F_J(t - t_{\min} - t_2 - \ldots - t_n)}{\mathbb{E}(J)}\right) g(x, \tau)\,\mathrm{d}x\,\mathrm{d}t\,\mathrm{d}\tau.$$

For other, fixed, turnaround times, $t_{\min}$ is replaced by the appropriate fixed time.

**Transmission probability.** To determine the transmission probability as a normalized force of infection $g(t, \tau)$ we used the infectiousness profile derivation of Ferretti *et al.* (*20*). We modified their derivation by noting that all environmental transmission can be traced back to an individual shedding into a shared environment at a particular time and therefore can be subsumed into infectiousness at that particular time. Ferretti *et al.* assumed that the infectiousness profile of an individual who develops symptoms is independent of the time of symptom onset, which assumes that symptom development does not reflect an immune response modulating infectiousness, that the symptoms themselves such as coughing do not increase infectiousness, and that the development of symptoms does not modulate potentially infectious behavior (a reasonable assumption very early in an epidemic, and perhaps increasingly reasonable as the duration of an epidemic increases beyond a few months). We note that this assumption may be weakened as further data become available. Ferretti *et al.* also assumed that the infectiousness profile of an individual who never develops symptoms is a linear multiple $\alpha$ of the profile of an individual who develops symptoms; again as more data become available this assumption may be weakened. Finally, they derived a Weibull distribution for generation time with shape and scale factors of 2.826 and 5.665, respectively. These assumptions amount, in our notation, to

$$g(t, \tau) \propto g(t),$$

$$\phi(1 - F_{M_P}(t))g(t) + \phi F_{M_P}(t)g(t) + (1 - \phi)\alpha g(t) = f_Y(t),$$

which simplifies to

$$\phi g(t) + (1 - \phi)\alpha g(t) = f_Y(t),$$

where $Y \sim \text{Weibull}(2.826, 5.665)$, and $g(t)$ is not required to satisfy the boundary condition $g(t, \tau) = 0,\ t > \tau$.

This framework takes into account the variability in incubation period for those who develop symptoms, but does not take into account the variability in the duration of shedding for those who always remain asymptomatic. We therefore modified the last equation by multiplying the second term by a factor of $(1 - F_{M_A}(t))$, finally yielding an equation

$$\phi g(t) + (1 - \phi)\alpha(1 - F_{M_A}(t))g(t) = f_Y(t).$$

We derived the distribution for $g(t)$ by solving this equation given the proportion of individuals who eventually develop symptoms $\phi$ and the relative infectiousness $\alpha$ of a completely asymptomatic individual versus an individual who goes on to develop symptoms. For $\phi$, we used the possible ranges derived above. For $\alpha$, we used the central value in Feretti *et al.* of an asymptomatic individual having one tenth the infectivity of a presymptomatic individual. We then set the relative infectivity of a paucisymptomatic individual between these two extremes, and derive $\alpha$ depending upon how the paucisymptomatic individuals are dichotomized between the asymptomatic and the presymptomatic groups. We then imposed our condition $g(t, \tau) = 0,\ t > \tau$, which is equivalent to assuming that all individuals with symptoms isolate. Finally, we normalized by $\int_0^\infty \int_0^\tau g(t) f_M(\tau)\, \mathrm{d}t\, \mathrm{d}\tau$ so that $g(t, \tau)$ satisfies the normalization condition.

Using this framework to estimate infectiousness for asymptomatic individuals leads to a small but in practice negligible discontinuity at the end of the infectious period.

The real-world infectiousness profile is influenced not only by disease progress in an individual, but also by that individual's behavior. Changes in population behavior have been shown to change the serial interval of SARS-CoV-2 infection (*21*). Of course, the aim of asymptomatic screening and isolation is itself to change the real-world infectiousness profile.

**Probability of detection.** The probability of detection is not simply the analytical sensitivity of the test being deployed. It is helpful to consider the components contributing to the probability of detection as consisting of ones independent from test to test (contributing to $p(t, \tau)$), and ones that remain the same from test to test (contributing to $p_\infty$). Some components will decompose into contributions to both $p(t, \tau)$ and $p_\infty$.

Components that remain the same from test to test broadly divide into factors that affect the probability of taking a test at all, and systematic factors that affect the sensitivity of the test but vary between individuals. For example, if the logistics of testing are arranged such that an individual can never access a test (e.g. occupational testing always performed on a day a worker is absent), the overall probability of detection in that individual is always zero. An individual who persistently refuses to be tested will be associated with an overall probability of detection of zero. An infected individual who always self-samples with an above average sampling technique will have a higher probability of detection than an otherwise comparable individual who always self-samples with a below average sampling technique. These aspects that do not vary from test to test contribute to the constant probability $p_\infty$ that an infected individual would *ever* be isolated following a positive test. This constant probability $p_\infty$ acts as an overall modifier to the proportion of transmission that can be abolished. (A simple illustration of this is to observe that if half of a population is never tested, then its contribution to the force of infection always persists.)

The probability that an individual is screened has substantial scope for variation, depending upon setting, ease of access to screening and attitude to screening, all of which correlate with demographic factors. Rate of uptake in established screening programs is known to be variable (*22*), and we should expect a variable rate of uptake in a SARS-CoV-2 screening program. For short screening programs aiming to test all staff in a hospital for SARS-CoV-2, uptake rates from 50–73% have been reported (*23, 24*), reflecting testing uptake in a population one might expect to be relatively motivated in a setting with relative ease for optimizing testing logistics. The longer term variability in individual uptake of SARS-CoV-2 tests is not well-documented in the literature, meaning we do not currently have a good appreciation of "screening fatigue" (how the uptake rate would change in longer term programs with regular repeat screening), and we do not know how to apportion reported rates of screening uptake between repeat-dependent ($p_\infty$) and repeat-independent ($p(t, \tau)$) parameters. We note, however, that all of these parameters would be easy to measure in any screening program where the size of the target population is known and it is possible to associate tests undertaken with individuals (to count the number of tests taken by each individual). In our realistic model, we set the probability that an individual ever attends screening (a contribution to $p_\infty$) to 0.8, and the probability that an individual who attends screening at least one attends on a given occasion (a contribution to $p(t, \tau)$) to 0.8.

The probability that virus is detected, usually referred to as test sensitivity, in fact has multiple components. Firstly, it is important to distinguish between analytical and clinical (or diagnostic) sensitivity (*25*). Many of the nucleic acid amplification tests (NAATs) used to detect SARS-CoV-2 have very high analytical sensitivities, reliably detecting a handful of copies of viral RNA in a sample. However, this does not necessarily mean that such a test has a high probability of detecting infection in an individual, since the probability of obtaining viral RNA in a sample may be significantly below 100%. Accurately estimating this clinical sensitivity is difficult owing to the lack of an appropriate gold standard comparator. It has been estimated to be between 46% and 98% (*8, 26, 27*). Factors affecting clinical sensitivity include anatomical sampling site and time during disease course of sampling. Expert opinion suggests that for reverse transcription polymerase chain reaction (RT-PCR) tests, 70% would be a reasonable working estimate of overall clinical sensitivity (*28*). Test sensitivity at a given time of sampling is likely to correlate with the amount of viral shedding.

We therefore modeled probability of virus detection by NAAT in two different ways. The first way uses the time-since-onset sensitivity profile reported by Zhao *et al.* (*8*). The advantage of this model is that it is based directly upon data relating detectability of viral RNA to other standards, rather than assuming a relationship between infectiousness and detectability. The disadvantage is that the time reference point used to anchor the detectability profile is onset of symptoms. This reference point is appropriate for making a clinical diagnosis but may not be most helpful for determining the relationship between infectiousness and probability of detection. Indeed, our model considers onset of symptoms to define when a person stops being infectious. To deal with this difference, we ran the model with this detection probability profile shifted by 3 days from the beginning of infectiousness. We added a linear interpolation of the detection probability from zero to the first detection probability reported by Zhao *et al.* The maximum sensitivity reported by Zhao *et al.* is 73.3%, meaning that, under this model, the contribution to $p_\infty$ can vary between 73.3% and 100%, with $p(t, \tau)$ adjusted accordingly so that the contribution to $p_\infty \cdot p(t, \tau)$ takes the values reported by Zhao *et al.* The second way we modeled probability of detection assumes the probability is related to the transmission probability $g(t, \tau)$. We modeled the probability of detection as being directly proportional to the transmission probability until the maximum transmission probability, then directly proportional to a timelag of the transmission probability, to reflect the persistence of detectable but non-viable viral RNA after the peak (*29–32*). We set the lag in our model at 2:1, i.e. the probability of detection at day $n$ after peak is based upon infectiousness at day $n/2$ after peak. This decision was based upon results of non-human experimental influenza infection (*33*). Direct evidence for the choice of lag in SARS-CoV-2 is currently limited. The scaling we applied sets the peak probability of detection to the maximum probability reported by Zhao *et al.* We emphasize the fundamental conceptual difference between these two methods of modeling the probability of detection, in that they use different time reference points to anchor the profiles: the method based on Zhao *et al.* assumes onset of symptoms is a reference point, whereas the method based on the transmission probability profile assumes that the onset of infectivity is a reference point. Where not otherwise stated, we calculated using the infectivity-based model rather than the model based on Zhao *et al.* We set the contribution to the sensitivity from $p_\infty$ to be 0.85. This makes the contributions to $p_\infty$ and the maximum value of $p(t, \tau)$ approximately equal.

Where the screening test is not a NAAT and has reduced analytical sensitivity, additional contributions to $p_\infty$ and $p(t, \tau)$ may be required. We considered a near-patient testing scenario using a preliminary report of test sensitivity of lateral flow devices (*9*). This report estimated that the assay technique had a sensitivity between 57.5% and 79.2% of that of NAAT, depending upon who administered the test.

There is one further modification to effective test sensitivity during screening not usually reported in the literature: the technical failure rate of the assay. All assays occasionally fail to detect evidence of infection because of a failure in the reaction: in the case of NAATs causes of such failure include the presence of inhibitory material in the sample preventing the reaction from progressing, and a technical failure in mixing reagents. Such failures are usually detected by failure of internal control material to amplify, reported as technical failures (i.e. no result), and not counted when considering test sensitivity. In the diagnostic laboratory an attempt at repeating the assay may be made, but in the high throughput setting it is possible that repetition would be infeasible. If a program were not to repeat testing in such cases, then $p(t, \tau)$ would need to be modified by a constant reflective of the technical failure rate. We have not included this modification in our model.

The final component contributing to our estimate for the probability of a successful detection is the probability that, following a laboratory detection of virus, an individual isolates and ceases to infect other people. In many settings, concordance with a request to isolate is unlikely to be absolute (one United Kingdom study reported intention to isolate in around 70%, and a rate of complete isolation at home amongst those who met criteria of 18.2% (*10*)), varying with cultural norms, demographic factors, incentives and enforcement (*34–36*). The consequent reduction in infectivity will vary; for the sake of our model we simplify this by considering a single, multiplicative contribution to $p_\infty$. Comparison with follow-up rates from historical screening studies with voluntary attendance at the screening stage suggests that a multiplicative contribution of 0.7–0.8 would be reasonable (*37, 38*); it may be possible to increase this multiplier with additional follow-up, incentives or enforcement. In our realistic model, we set the probability that an individual who screens positive isolates to 0.7.

We investigated the effect of varying individually these components contributing to the probability of detection. To generate an estimate of the real-world effect of such screening, we set $p_\infty$ equal to 0.476 (the product of probabilities of 0.8 for ever attending screening, 0.85 for invariant screening sensitivity, and 0.7 for isolation). We assumed a contribution to $p(t, \tau)$ of 0.8 arising from the per-time probability of attending screening amongst those who ever engage. We further assumed a multiplicative contribution to $p(t, \tau)$ from the probability of detection of the relevant value from reference (*8*) divided by 0.85. To generate an estimate of an idealistic scenario for comparison, we modeled the situation where all individuals attend screening and isolate perfectly, i.e. where $p_\infty$=0.85 and $p(t, \tau)$ is the relevant value from reference (*8*) divided by 0.85. This allows comparison between the technical capabilities of screening and the anticipated effects of screening with real-life behavior taken into account, to allow determination of the feasibility of a screening program with behavioral interventions.

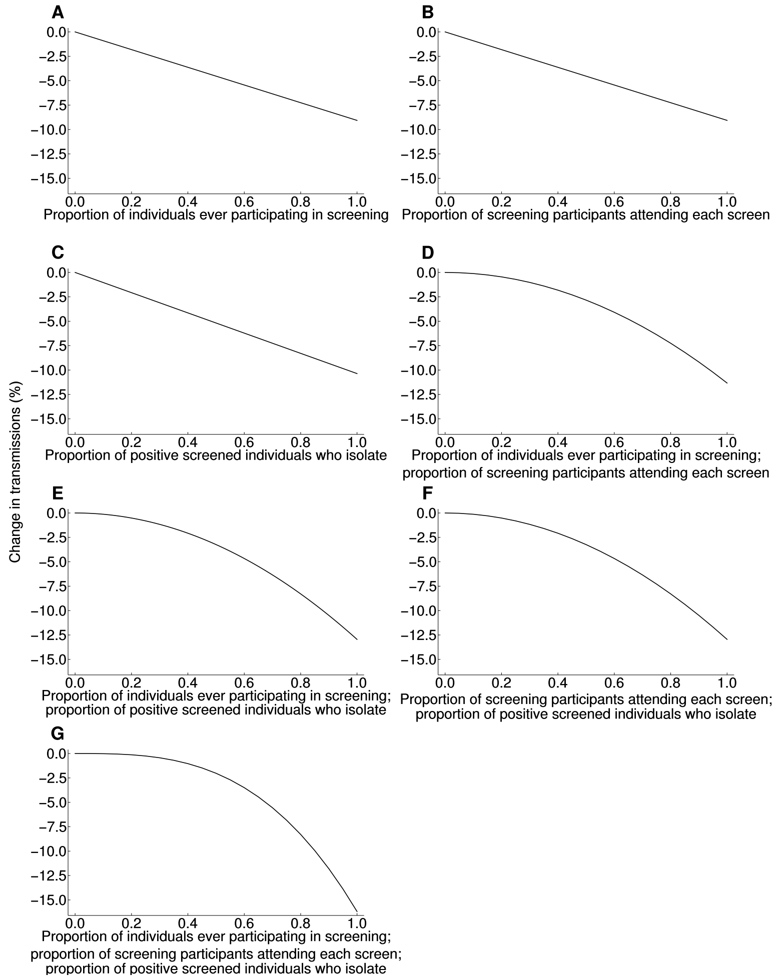


**Fig. S1. Effect of changing engagement with screening on transmissions.**
For the realistic testing scenario, offering weekly screens. Where not varied, the proportion of individuals ever participating in screening is set to 0.8, the proportion of screening participants attending each screen to 0.8, and the proportion of positive screened individuals who isolate to 0.7. The contribution of each of these proportions to transmission is shown to be linear or approximately linear. The contributions can multiply together. (**A**) Effect of varying proportion of individuals ever participating in screening alone. (**B**) Effect of varying proportion of those who ever engage in screening that participate in a given screen alone. (**C**) Effect of varying the proportion of positive screened individuals who isolate alone. (**D**) Effect of covarying proportion of individuals ever participating in screening with proportion amongst those individuals who engage in a given screen, using identical proportions for each. (**E**) Effect of covarying proportion of individuals ever participating in screening with proportion of positive screened individuals who isolate, using identical proportions for each. (**F**) Effect of covarying proportion of those who ever engage in screening who participate in a given screen with proportion of positive screened individuals who isolate, using identical proportions for each. (**G**) Effect of covarying all three proportions together, using identical proportions for each.

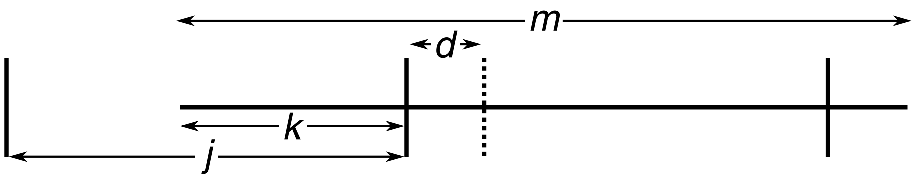


**Fig. S2. Schematic representation of the screening scenario.**
Time is represented horizontally. Asymptomatic/presymptomatic virus shedding lasts for duration $m$, starting $k$ time before the screening event (solid vertical lines) in an interval between events of length $j$. There is a turnaround time $d$ before a positive result is acted upon (dotted vertical line).

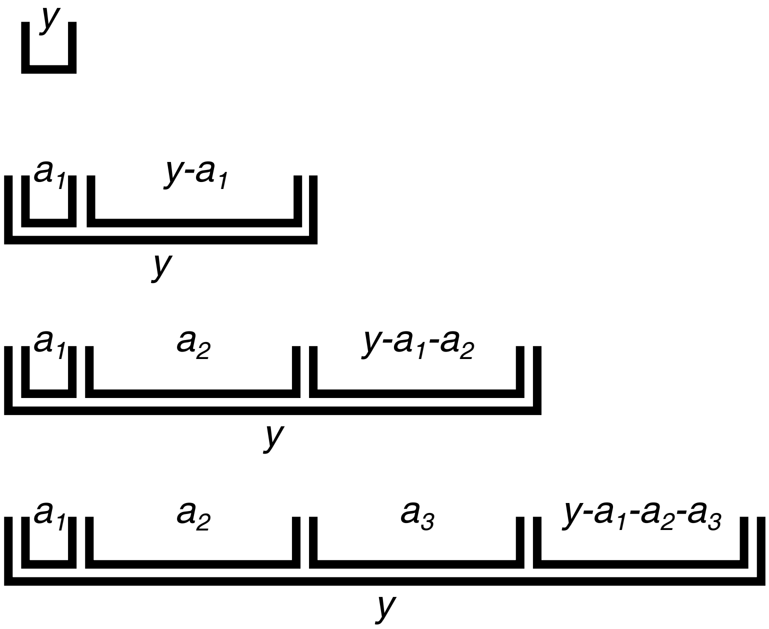


**Fig. S3. Schematic representation of repeat screening.**
Time is represented horizontally from the onset of viral shedding. Successive rows represent the construction of the convolutions for a successful screen after one, two, etc. attempts. In this example, for ease of illustration, the time between successive screens is represented as being constant and identical, but it may in principle vary.

| Scenario | Predicted transmission reduction (%) |
|---|---|
| Our realistic scenario | 7.25352 |
| Shedding: Xia *et al.* (*15*) raw data | 7.20066 |
| Shedding: Xia *et al.* (*15*) derived Weibull distribution | 7.21595 |
| Shedding: Lauer *et al.* (*16*) lognormal distribution | 7.16608 |
| Transmitting: asymptomatic infectiousness 0.01 rather than 0.1 | 7.39690 |
| Transmitting: asymptomatic infectiousness 0.5 rather than 0.1 | 6.99344 |
| Transmitting: asymptomatic infectiousness 1 rather than 0.1 | 6.69431 |
| Transmitting: asymptomatic infectiousness 2 rather than 0.1 | 6.38360 |
| Transmitting: asymptomatic infectiousness 10 rather than 0.1 | 6.30979 |
| Detection: Zhao *et al.* (*8*) detection profile | 7.26128 |

**Table S1. Effect of changing model assumptions on predicted transmissions in the realistic scenario.**
In each case the predicted transmission reduction is listed for a scenario that is the same as our realistic scenario except for the one change listed. The scenarios and all assumptions are described in detail in the Materials and Methods. Given that this table aims to illustrate limitations in model accuracy arising from parameter choices, results are reported beyond the expected accuracy of the model, to enable comparison.

**Caption for Code S1.** Mathematica code implementing the screening models to produce the results in this paper.
